# Understanding maternity care providers’ use of data: A qualitative study in Southern Tanzania

**DOI:** 10.1101/2022.10.11.22280938

**Authors:** Regine Unkels, Fadhlun Alwy Al-beity, Zamoyoni Julius, Elibariki Mkumbo, Andrea B Pembe, Claudia Hanson, Helle Mölsted-Alvesson

## Abstract

**Introduction:** Health information management system data is collected for national planning and evaluation but rarely used for health care improvements at the sub-national or facility-level in low-and-middle-income countries. Research suggests that perceived data quality and lack of feedback are contributing factors. We aimed to understand maternity care providers’ perceptions of data and how they use them, with a view to co-design interventions to improve data quality and use.

**Methods:** We based our research on constructivist grounded theory. We conducted 14 in-depth interviews, two focus group discussions with maternity care providers and 48 hours of observations in maternity wards of two rural hospitals in Southern Tanzania. Constant comparative data analysis was applied to develop initial and focused codes, sub-categories and categories continuously validated through peer and member checks.

**Results:** Maternity care providers appropriated numeric data on service provision to reconcile their professional values and demands from managers and the community with effects of a challenging working environment. They felt controlled by their managers’ data requirements and alienated from service provision data. Providers added informal documentation ways for their own narrative data needs to reflect on and improve service quality. These also assisted them to recreate social relationships with managers, clients and the community. The resulting documentation system led to duplication and increased systemic complexity.

**Conclusions:** Data from health information systems does not represent an independent and neutral entity but is embedded into the social realities of different users. Appropriation and use of data reflect these realities and users’ working environment. Interventions to improve data quality and use may need to incorporate the multitude of clinical and administrative documentation and data needs to avoid duplication and inefficiencies.

## INTRODUCTION

Data on service provision from health facilities is considered vital for health system strengthening. Data captured by *Health Management Information Systems* (HMIS) informs national and sub-national health system performance monitoring including i) monitoring and evaluation, ii) resource planning and iii) service management (1). These purposes underlining the numeric nature of health information system data apply globally, although high-income countries mainly rely on electronic registries collecting individual patient data rather than aggregated facility data (2). In most low-and middle-income countries (LMIC) like Tanzania, the HMIS is organized around pre-printed registers and forms filled by health care providers. Monthly summaries are later manually digitized into the *District Health Information System* (DHIS2) (3, 4). Additional official forms of documentation are used at the facility-level i) to support data generation for HMIS such as admission- and discharge-registers or ii) for decision-making, such as the partograph, but research on HMIS rarely include these (5, 6).

Studies frequently describe problems with data quality for HMIS. Commonly cited findings are incorrect or incomplete recording in primary registers and a mismatch between registers, reporting forms and DHIS2 data (7-10). The studies suggest that these issues can be improved through increased supervision and feedback at facility level (11). Underlying reasons for low data quality at health care provider level are, however, poorly understood (12). Moreover, it has been suggested that data quality and data use are linked (13, 14). This is underscored by emerging evidence that health facility data is rarely used at district and facility level (15, 16). Apart from perceived low data quality, other factors explaining the limited use were lack of i) feedback, ii) accountability for data quality, iii) information-use culture (17-20) and the HMIS’s unresponsiveness to shifting data demands and priorities (2). The importance of an organizational culture emphasizing data-informed decision-making within health systems, coined as *information-* or *data-use culture* is increasingly propagated to improve accountability and data use in LMIC (21, 22).

The problems described raise concerns about the effectiveness of HMIS as the most suitable data collection system (2, 23). HMIS data collection processes and low data use may lead to a situation where clinical staff, such as maternity care providers (MCPs), perceive themselves as mere data producers, failing to use data for clinical care purposes (4, 5).

The aim of this study was to improve understanding about data needs of maternity care providers in Southern Tanzania and which type of information is used together with HMIS data for which purpose.

## METHODS

We conducted this qualitative study with MCPs of two hospitals in Southern Tanzania. The *Consolidated Criteria for Reporting Qualitative Studies* (COREQ) is used to report here (24).

### Setting

We included one district and one regional hospital. The hospitals served a poor rural population living of subsistence farming (25). District hospitals in Tanzania, with 100 to 175 beds, typically provide antenatal and postnatal care, routine labour and emergency obstetric care. Nurses and midwives at certificate or diploma level and non-physician clinicians or medical doctors work in maternity care. Regional hospitals have 176 – 450 beds and offer all the above with additional specialist care (26). Like other hospitals in Tanzania, the included hospitals faced important human resource challenges with approximately half of the required nursing and clinical staff available (26).

In Tanzania, HMIS documentation for maternity care includes three paper-based registers for i) antenatal care, ii) labour and delivery and iii) postnatal care, daily tally sheets and monthly summary forms. Since 2013 this data is subsequently digitized into DHIS2. This procedure was followed in the included district hospital. The regional hospital used an additional, locally developed electronic health information system collecting clinical and managerial information. Typically, health care providers receive infrequent training on HMIS. Recurrent re-allocation of staff contributes to attrition of knowledgeable staff (27).

Hospital management had introduced supplementary documentation e.g. admission, referral and discharge registers. The partograph, recommended for labour monitoring by World Health Organization (WHO), was integrated in pre-printed clinical patient files. These documents supported completion of patients’ antenatal care cards, HMIS tally sheets and summary forms.

In 2015 the Tanzanian government introduced an electronic *hospital management information system* (GoTHOMIS) with interfaces to existing digital systems like DHIS2 (28). This system was not functional in maternity wards during the time of data collection (table 1).

**Table 1:** Main systems for collecting service provision data in maternity wards of included hospitals

| Name of System | Abbreviation | Components | Official purpose | Introduced by |
| --- | --- | --- | --- | --- |
| Health Information Management System | HMIS | Printed register, daily tally sheets, monthly report forms<br>DHIS2 software | Health system planning | Ministry of Health |
| Government of Tanzania Hospital Management Information System | GoTHOMIS | Electronic registry | Health system planning | President's Office - Regional Administration and Local Government |
| Antenatal Care Card | ANC card | Printed card | Clinical documentation for decision-making | Ministry of Health |
| Partograph | - | Form integrated in clinical patient file | Clinical labour monitoring, decision-making, number of completed partographs entered in monthly report form | Ministry of Health |
| Electronic Information System (locally developed for one included hospital) | EIS | Electronic registry | Hospital management & planning | Hospital management |
| Clinical patient file | - | Locally printed form (admission, labour monitoring, delivery, postpartum care until discharge)<br>Observation charts | Clinical documentation, support to HMIS documentation | Commissioned by Ministry of Health, Hospital management |
| Admission book | - | Hand-written register | Support to HMIS documentation | Hospital management |
| Discharge book | - | Hand-written register | Support to HMIS documentation | Hospital management |
| Informal documentation | - | Referral book<br>Ward round book<br>Maternal /newborn death register<br>Shift report book<br>Work plan (district hospital)<br>Theatre/Caesarean section register<br>Equipment/supplies register (District hospital)<br>Emergency drug register (Regional hospital)<br>Loose paper notes | Various unofficial purposes | Nurse in charge of maternity together with maternity care providers |

### Study design

We used a qualitative study design, based on constructivist grounded theory (30, 31), with i) in-depth interviews (IDIs), ii) observations in maternity wards and iii) focus group discussions (FGDs) (supplementary figure 1).

### Sampling, recruitment, data collection and analysis

We report on sampling, data collection and analysis together in line with grounded theory (30, 31). Included hospitals were selected based on an heterogeneity assessment to ensure representation of hospitals in rural Tanzania (32). The first data collection with 14 IDIs and 48 hours of observations took place in February 2021. The second data collection, two FGDs (11 participants), was conducted in June 2021. MCPs working in maternity ward were eligible. We used theoretical sampling (30) to include a variety of cadres with experience in service documentation. Sampling for observations included all staff working in maternity at the time of observation (table 2).

**Table 2:**
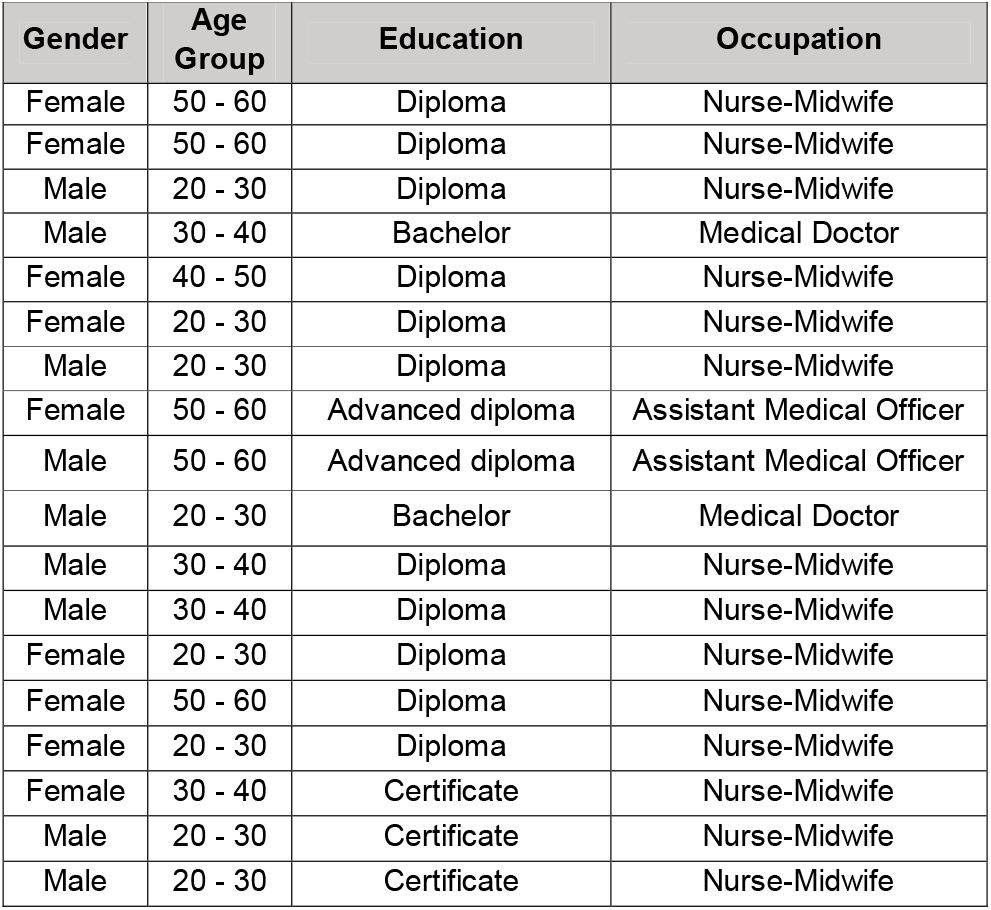
Demographic details of participants

MCPs received written and verbal information during initial encounters and could ask questions. IDIs, observations and FGDs were held at subsequent visits. We evaluated saturation continuously through simultaneous data collection and analysis to determine emergence of new information or topics.

Topic guides were grounded in previous research on HMIS data collection in Tanzania (4) and included topics like i) data use and usefulness, ii) documentation tools for different purposes and iii) views on data quality requirements among others. They were developed in English, translated into Kiswahili and pretested. Topics for the FGDs were derived from continuous data analysis. We used diagramming adapted from *user journey mapping* (33, 34) during FGDs to visualize MCPs’ data encounters (35) (figure 2).

**Figure 1:**
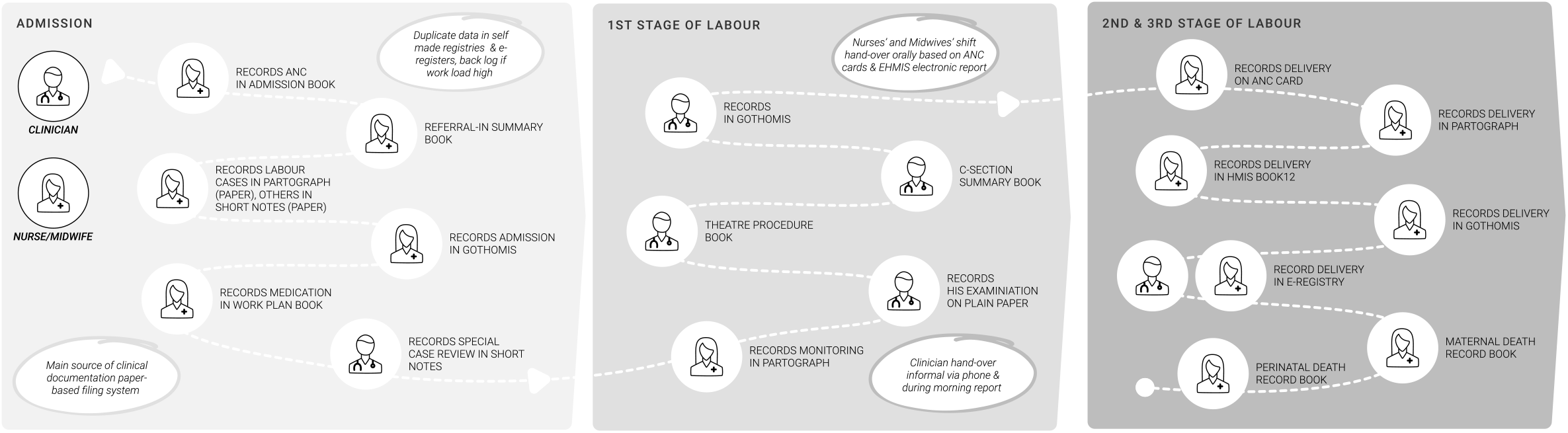
Study design.

**Figure 2.**
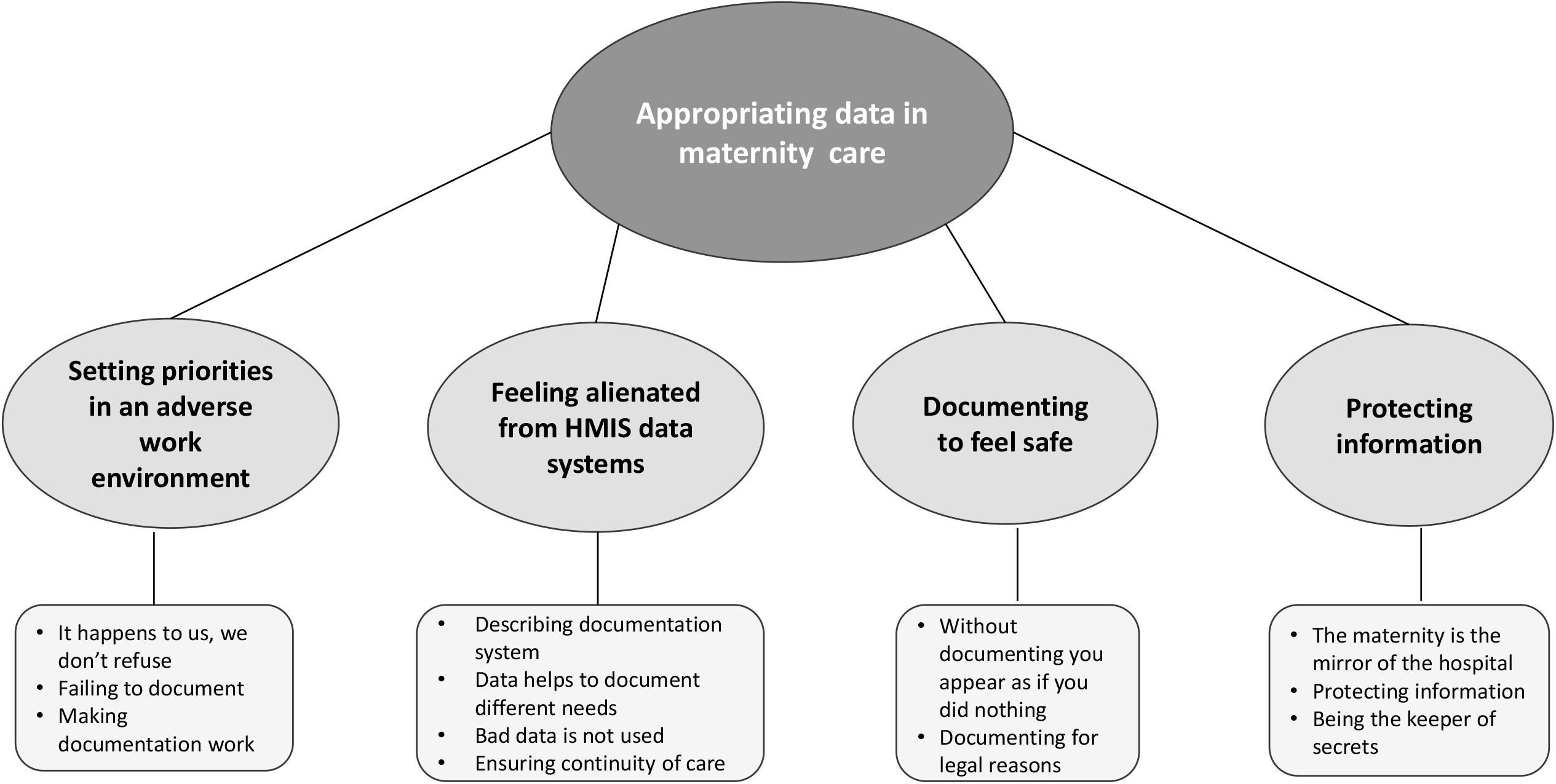
supplement: Example diagramming.

Observations took place at different times during daytime and night. IDIs and FGDs were held in separate hospital rooms with adequate audio-visual privacy. Interviews and FGDs were recorded and lasted one to two hours. Participants received refreshments and transport compensation from home.

Audios were transcribed verbatim in Kiswahili. Relevant quotes were translated into English. Observation notes and memos were taken in English. RU and FAB conducted initial coding on four transcripts in NVivo (NVivo v12, QSR) for preliminary analysis. Parallel analysis during data collection was also guided by memo writing and daily debriefing with all data collectors. RU performed initial line-by-line coding and subsequent focused and theoretical coding (30) on the complete data set. Constant comparative analysis was used to explore differences between i) individual transcripts, ii) data collection methods and iii) hospitals. Codes, categories and memos were continuously reflected for theory building through frequent peer check with FAA, HMA, EM and ZJ. We conducted member checks during FGDs as part of the ALERT co-design process and at category level (11 MCPs from four ALERT hospitals), to enhance trustworthiness (36, 37).

### Patient and public involvement

We did not involve patients or the public in this research due to the subject matter. They were not invited to contribute to design, analysis nor manuscript review.

### Reflexivity

The research team included early career researchers (RU, ZJ) and experienced researchers (FAA, EM, ABP, CH and HMA) from middle- and high-income country institutions. RU, FAA, ZJ, ABP, CH have medical backgrounds. HMA and EM are medical anthropologist, social scientist. Three co-authors are male and four females.

FAA, RU and EM participated in data collection for IDIs and observational data with one social scientist and two nurses (all male) experienced in qualitative research. RU and ZJ collected FGD data. All had worked in the study area and speak fluent Kiswahili. The research team maintained an open conversation throughout data collection and analysis. All members have access to the data.

Senior writers (ABP, CH and HMA) supported less experienced researchers through an open process manuscript revision. Findings of this research will be integrated in the ALERT dissemination through open access publication and during various stakeholder workshops. Our findings will contribute to the Tanzanian government’s endeavor to improve routine health data collection.

### Ethical considerations

Clearance was obtained in Tanzania from the Institutional Ethics Review Board of *Muhimbili University of Health and Allied Sciences* (MUHAS-REC-4-2020-118), from the *National Institute for Medical Research* (NIMR/HQ/R.8a/Vol IX/3493) and from the *Swedish Ethical Review Authority* (2020-01587). Written consent from participants and written institutional consent for observations at maternity wards as well as assent from MCPs, were obtained.

## RESULTS

We report results based on four categories and 12 sub-categories, linking to one core category (figure 3).

**Figure 3:**
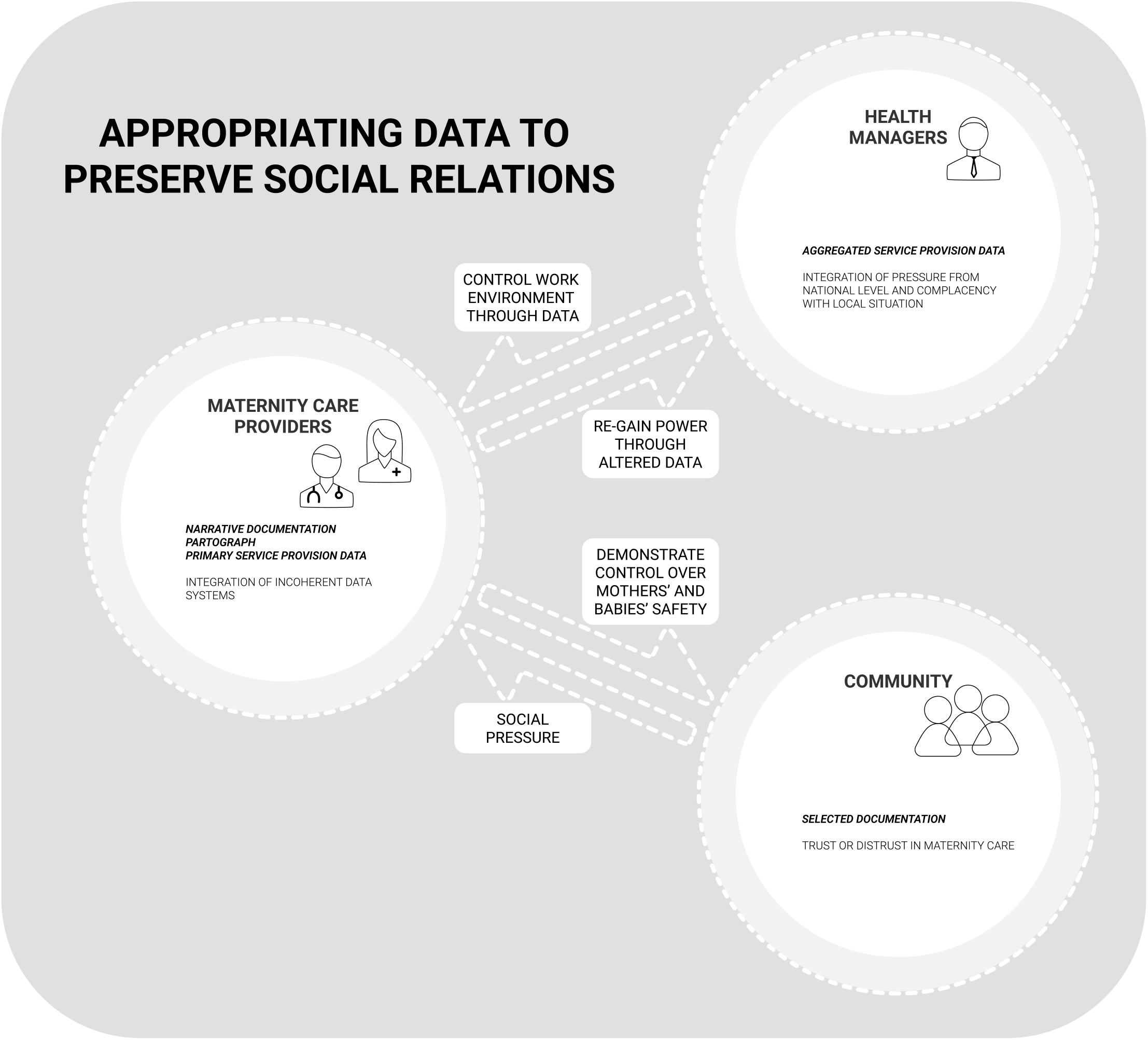
Core category, categories and sub-categories.

Our analysis resulted in a theoretical model that depicts how MCPs appropriate official HMIS data and informal data sources to preserve social relationships with various stakeholders of maternity care (figure 4).

**Figure 4:** Visual theory representation.

### Category 1: Setting priorities in an adverse work environment

Participants described their difficult working environment and how this affected their decision-making regarding task prioritization. Challenges included i) low staffing levels, ii) high patient numbers and iii) occasionally, missing documentation tools, foremost hard copies of the partograph or antenatal care cards. MCPs argued, they could not conduct documentation and care concurrently when number of clients and allocated tasks clashed.

During observations it was noted that fetal heart rate (FHR) was measured infrequently, although partographs showed documentation of measurements every 30 minutes. MCPs assigned a high theoretical value to the partograph emphasizing that they did understand the importance of measuring FHR according to standards, and how they felt when they did not live up to this.

> “*30 minutes have passed I should quickly listen [to the fetal heart rate] again. And when I lose that time, I feel bad because it is not my intention to do so but the work is overwhelming*.” (Interview Nurse-Midwife, female)

Participants explained that, with available tools and few patients, documentation was easy. In contrast, if they had to care for emergencies or many patients, MCPs limited documentation to the patient’s antenatal care card only. Other documentation would then be done at the end of a shift together.

> “*For example, here we are two per shift but there are times when every bed is occupied. Then it is difficult: Once you come out here, you go to this patient, then to this one. For a long time, you move around. Until you come back to the first one may take some time*.” (Interview Nurse-Midwife, female)

### Category 2: Feeling alienated from HMIS data

All MCP knew, completing the HMIS was their task, although some would have preferred to employ data clerks for this. MCPs and immediate managers had created other documentation types to maintain reporting when HMIS tools, e.g., registers, were lacking, or as a back-up if electronic data collection systems were unavailable. They had also added separate registers to document deaths or for shift reporting. The same information was thus recorded in several documents since official registers still had to be completed once available.

> *“You must fill more than one register, even more than two. You start with the partograph, then two registers. So, the time you spent on this is more than the care you provide”*. (Interview Nurse Midwife, male)

Apart from duplicate work this sometimes resulted in incongruent data between different documentation types.

Participants described themselves as mere data producers, only receiving feedback from managers on their data when things went wrong and reported not to use this data for their daily work.

> “*For now, I don’t see anything [in terms of use]. We collect the information, we submit it, like there are 200 women who delivered normally, five had a Caesar [Caesarean Section]. That’s it, you have already left it with the bosses, they have taken it and left with it*.” (Interview, Nurse-Midwife, male)

In addition, HMIS data was perceived as a managerial means to control MCPs’ performance or workload. Participants described using HMIS data to prove their hard work and, ultimately, to justify their employment in maternity.

> *“…. Because I as a midwife if I say we have [many] deliveries, I must know how many, maybe it is 200, but if I don’t have data, it is like there are zero*.” (FGD, female)
>
> *“Yeah, you know, it is important to do this [documentation] because without documentation you are perceived…. as if on that day you haven’t worked, you just sat there*.*”* (Interview Nurse-Midwife, female)

Accountability for data was seen as conditional to an enabling environment created by management.

> “*My perspective concerning accountability is first we should have a friendly environment that will make everyone see themselves as the person responsible for completing the information*.” (Interview Nurse-Midwife, male)

MCP mentioned immediate usefulness as the main determinant of what constitutes good data. Participants described data e.g., as good if they solved problems.

> “… *Bad data is the one that you just enjoy, you take it but don’t use it*…..*Good data is sustainable, meaning that you can collect this data then you go and use it, and it solves the problems that exist somewhere*.” (Interview, Assistant Medical Officer, male)

They described how other ways of documentation satisfied their immediate reporting needs, e.g., for communication about patient care, especially during shift hand-over. New informal documentation tools were often added either by the nurse-in-charge or MCPs to improve documentation after an incident, e.g., a newborn death.

> *“We mentioned that we have this notebook here, that documents when we get a patient back [from theater], when we transfer her [to the ward] and who receives her. The purpose is, if there is a gap anywhere, if we delay somewhere, so that we got an unfavorable result, that we can use this to evaluate so that we can improve*.*”* (Interview Nurse-Midwife, female)

Most of these formats were informal narratives, and not necessarily reviewed by managers (table 1). They consisted of i) paper notes, e.g., from clinicians, who didn’t document on the partograph, ii) hand-written observation notes for shift hand-over, iii) little notebooks, e.g., to document patient hand-over between theatre and maternity or iv) patient files where cases were classified according to severity. A very important piece of added documentation in one hospital was a register to document medication and during observation midwives were seen reading it frequently or discussing it.

> *“…For those women with regular i*.*v. medication we have developed a work plan register that shows e*.*g*., *a mother receives powercef at this time, crystapen at that time*
>
> *…You know, this really helps us not to forget to administer medication. So, this “that patient, I have forgotten”, that doesn’t occur*…*”*. (Interview Nurse-Midwife, female)

### Category 3: Documenting to feel safe

The partograph was portrayed as the centerpiece of MCPs’ documentation efforts for this purpose. Nurses talked about the importance of using the partograph and archiving it to access it when mothers came back with a sick newborn. Partographs were kept together in piles in cupboards after they had been counted for the monthly HMIS report, but participants mused about the need to produce something in writing to reduce problems with the community they were so close to.

> “*It can happen that she comes back for other problems before the end of those 42 days*… *So, if she comes you should show it [the partograph]. Maybe the baby has a problem*.*”* (Interview Nurse-Midwife, male)

Apart from social risks of bad outcomes, like being blamed by the community, participants were also afraid of legal risks and written documentation seemed to help them cope with this.

> *“Yes, we all check [fetal heart rate], I count and write and the one who takes over from me also does that. So tomorrow, there is documentation showing that at 1:40 I have checked, and I have left [the patient] with you and from then on you have checked. And*… *[the document] will show what you did until that fresh stillbirth happened. And if the document doesn’t exist, we will lock ourselves up [in jail]*.*”* (Interview Nurse-Midwife, female)

Supervisors were aware of the challenges with partograph completion. They urged staff to fill incomplete partographs retrospectively, because the number of complete partographs was included into monthly reporting to higher level.

> *“…because the supervisor is here so if you don’t fill, she will know and say: “look at that, here you haven’t completed, complete this because if you don’t, we will have a gap when we report [at the end of the month] and we will suffer*.” (Interview Nurse-Midwife, male)

### Category 4: Protecting information

Participants argued that service provision data was hospital property and ensured that the information they generated was protected from the view of others, i.e., relatives and patients. Their documentation of provided care, or rather written scenarios of their perception of that care, should not be shared with women and their companions.

> “*The mother cannot go [home] with the partograph since it is our property. There is a lot of confidential information written, so we keep them in our files which stay in a cupboard*.” (Interview Assistant Medical Officer, female)

MCPs explained their role as keepers of secrets, their own and others’, to preserve social integrity for the community to which they belonged.

> “*Hhmm, the most important is confidentiality because the people we serve, are the ones we are meeting in the streets. It is the community that surrounds us. So, the most important way to protect medical information is confidentiality*.*”* (Interview Asisant medical Officer, female)
>
> “… *At the end of the day the news will spread, …The woman in question will hear or other people will hear, so if this happens, it is not good, we may lose clients, mothers will be afraid to come… Some have serious problems, so it is very important to keep their secrets*. (Interview Nurse-Midwife, female)

They cited confidentiality as one reason why companions couldn’t be allowed inside during childbirth. On the other hand, the latter were often asked to take clients’ antenatal cards for registration even though it contained confidential information, e.g., a mother’s HIV status.

### Core category: Appropriating data in maternity care

We identified MCPs’ appropriation of data, to create a more desirable social narrative for themselves, their supervisors, and the community, as the core category for theory development (figure 4).

MCPs explained how i) their own professional values, ii) their need for feeling safe during work and iii) the importance of maternity services for the community, often collided with their working environment and managerial pressure.

To make sense of these situations MCPs changed the meaning of health service data from the official health system perspective, i.e., for the use of someone else, to a personal perception, where data provided them with a sense of control over an environment that was perceived as adverse, the community’s opinion about maternity care and providers, and over managerial regulation of their work performance.

MCPs tried to uphold a positive image of their professional self and of the care provided, for their own sake and for the community, despite the challenges they were facing during their work. This is summarized by the quote below:

> “*We are a small number [of MCPs] but together we have decided that despite being so few we must document the things we do. It is important, because this is maternity, it is the mirror of the hospital*.” (Interview Nurse-Midwife, female)

Altered FHR data for example, could show that monitoring of fetal and maternal wellbeing was done according to standards and that ultimately good care was delivered.

Participants repeatedly emphasized the importance of documentation in general to underline their trustworthiness and to rebut managerial control. Altered HMIS data could also potentially conceal a situation, where more staff was available for fewer deliveries. During observation, we noted that indeed shifts were not always busy. Participants rarely mentioned these situations though, but rather described scenarios where either too many laboring women or women with serious complications met with too few staff.

Altered and appropriated partograph data assisted MCPs to feel safe when care went wrong, but other official data sources did not fulfill this need. Additional documentation sources were therefore created to ensure that communication and documentation supported MCPs in case problems with suboptimal care occurred. Immediate managers were complacent about alteration of partographs to meet their own requirements, thus facilitating data modification.

This complex situation of added documentation and appropriation of HMIS data for social purposes led to little accountability towards official data systems, their purposes and documentation in general.

Data could be altered or presented in a different way to safeguard social relationships, but it could also be withheld for the same purpose. MCPs and their managers closely lived within the community they provided care for, making them vulnerable in the context of their adverse working environment. Protecting their own documentation seemed thus important to reduce social disruption through breaches in confidentiality. On the other hand, MCPs could choose whether to hold back or share their clients’ sensitive health data, such as a positive HIV-status, which may have contributed to their stand in the community.

## DISCUSSION

In summary our results suggest that MCPs integrated formal and informal data on service provision to unite diverging influences from working environment, management, society, and their professional perception into an alternative account of maternity care. HMIS data, the partograph and narrative documentation together embodied these different relationships (figure 4). This enabled MCPs to feel protected against litigation, managerial mitigation measures and against disrupted social relationships with the community. MCPs felt alienated from numeric HMIS data prioritized by managers, because it did not fulfill their need for narrative data. To satisfy this, they added hand-written records, and consequently another layer of complexity to the already intricate HMIS. Health care managers were complacent with data modification to meet their own data requirements. This led to low individual and systemic accountability towards quality and use of service provision data.

### Core category: Appropriating data in maternity care

The concept of social dimensions of data and its use to construct realities of self, work performance or health has gained much attention with increasing digitalization of human life and interactions (38, 39). Lupton describes how a seemingly objective, quantifiable entity like personal data was used by lay people to make sense of their body and gain control over their health in the context of self-tracking devices (40, 41). Research suggests that numeric data is not only embedded in social interactions but can also embody them. This notion has been applied to health service provision data regarding caesarean section in the United States and maternal mortality in Malawi (42, 43). Wendland depicts the apparent divide between seemingly neutral numeric indicators on one hand and how this evidence is contextually shaped on the other. Other authors report, how numeric data is modified by health care providers to counteract pressure from health management in Burkina Faso, Ethiopia and Tanzania (5, 6, 44). Our participants illustrated how HMIS data was adapted, and narrative documentation tweaked to construct a seemingly more trustworthy reality, with perceived control over work environment, managers’ and public reactions to their performance. Previous research by our group from Southern Tanzania also describes MCPs’ accounts on how data in official HMIS registers is altered to satisfy official requirements (4).

Estifanos et al. take a health system focus and emphasize on managerial pressure as a main driver for data modification in Ethiopia (44), but we argue, that health care data is part of a more complex social, organizational, and individual concept of power, alienation, accountability and social integrity (figure 4).

### Category 1 Setting priorities in an adverse work environment

Our findings depict how a challenging work environment shaped MCPs’ appropriation of data to align professional, health system and societal priorities. Others have also described the environment of maternity care and documentation in Tanzania as volatile (45) and complex (6), often constrained by a lack of human resources and commodities (46).

Some authors imply that organizational culture may inform MCPs’ view of their tasks. Research on the use of HMIS data including our own, suggests that MCPs need role models and support to develop a data perception that is led by accountability and transparency (4, 8, 22). In line with our findings, MCPs in Burkina Faso and Ethiopia reported managerial complacency with data modification (5, 44). These experiences are likely to shape an organizational culture where data is used to safeguard social relationships as a coping mechanism within a complex environment.

### Category 2: Feeling alienated from HMIS data systems

Most research on health data focusses on official HMIS registers only (47, 48). Much less attention has been given to MCPs’ efforts to increase use of data for clinical care, through added documentation: Strong describes how MCPs in Tanzania interacted with numeric data for hospital and health system purposes, where MCPs entered information into a panacea of notebooks and registers increasing documentation load (6). Our results support this finding but add a new perspective by illustrating how MCPs used added documentation to fit their own data needs.

We describe how MCPs felt alienated from the official data: They could not use it immediately and only received negative feedback on their efforts. The data was thus perceived as useless and consequently accountability was low. Other research from similar settings supports these findings: Numeric data was seen as owned by the management (5, 44) and, similarly to our results, its completeness was valued over correctness (12). MCPs saw themselves as producers of data rather than users, feeling disempowered to apply their own data (4). Our participants described actual use of their narrative data while participants from our previous research reported that data which could be useful to them was missing from HMIS (4).

This finding introduces the question how applicable numeric service provision data is to quality improvement at health facility level. The main purpose of HMIS is to generate coverage data for the purpose of planning and performance monitoring (2, 49), thus focusing on the managerial part of health care. These systems developed over centuries and with an almost organic growth rather than design, user perspectives, like our participants’ views, were often neglected (50) in the past. In our study, MCPs and managers added other narrative dimensions to the official system, to tie incoherent purposes for different users. This process resulted in making the documentation system even more complex.

### Category 3: Documenting to feel safe

Our participants reported how they used documentation to feel safe in a work environment where good birthing outcomes were not guaranteed. These reports are in line with other research from similar settings where MCPs used altered partographs to protect themselves against managerial reprimand action or legal consequences in cases of maternal deaths (5, 6).

WHO has promoted the partograph for labour monitoring since long although effects on maternal and fetal outcomes were not ascertained (51). MCPs in our study worked in an environment that often prevented them from meeting official standards but they still claimed that the partograph was their most important document. This claim enabled them to uphold a core professional value of maternity care: To make sure mothers and babies are safe, despite different realities in their maternities. Participants argued that the “*maternity ward was the mirror of the hospital*”, i.e. hospital performance was judged by their work. Our participants used altered partographs to create a new narrative of a maternity care where standards were followed, albeit on paper. MCPs reasoned that this information should be kept well because when mothers came back with a sick newborn, they had to demonstrate that this child had been doing well under their care. These reflections may be interpreted as their attempt to feel safe in this aspect, despite the circumstances, as in reality it was quite difficult if not impossible, to retrieve an individual partograph after discharge.

### Category 4: Protecting information

Participants described the importance of confidentiality for their acceptance within the community. They explained that confidentiality issues may prevent clients from coming to the maternity and other research suggests that fear of disclosure may deter HIV-positive women from accessing services (52). We hypothesize that MCPs power on decisions whose and which data was shared, may have added to their social standing.

Our results add new insights to the literature on health information systems and data use: How data and documentation may reflect the provider-client-relationship. In rural, remote areas such as our research setting, MCPs usually live within the small communities they serve, with reciprocal interdependencies. The balance between integration into a rural community and rejection may be delicate (53). This may explain our participants’ need to protect social integrity, and to include their clients in the creation of an alternative narrative of maternity care (figure 4).

### Methodological considerations

The strength of our study is the use of a variety of qualitative methods for triangulation, including member and peer check at different analysis levels. Diagramming helped participants to reflect on the different types of data they produced and to describe their complex system of documentation to the researchers. Some of the data collectors were known to participants including the European lead author. This may have led to social desirability bias. We used triangulation and frequent member/peer check to mitigate this potential bias.

We prioritized health personnel to gain deeper insights into their data use without including the opinion of health care managers or community members. Managers’ voices have been captured in a previous research study (4) and since MCPs generate the primary data, it deemed important to further understand their perceptions and narratives.

Only four out of 18 participants were clinicians (two medical doctors and two assistant medical officers). This reflects the professional distribution in included maternities.

## CONCLUSIONS

Health data is not neutral, and users assign different meanings to it. Policy makers need to see health information systems as one component of the wider health system where other parts, such as staff and supply availability, should be improved to influence MCPs realities which may then affect data quality and use.

Current health information systems may not reflect maternity care providers’, or other healthcare staffs’, data needs. These systems have not been designed but have rather evolved over time and relevant narrative parts for clinical uses are missing. They should be re-designed with an “all-data-view” in mind, taking all users on board, including MCPs and their clinically oriented narrative data.

## Supporting information

Supplementary COREQ checklist

## Data Availability

All relevant data are within the manuscript and its Supporting Information files. Primary data such as transcripts reflect views of maternity care providers from two health care facilities in Southern Tanzania, with a very small population of maternity care providers. Making the full data set publicly available could potentially breach the privacy that participants were promised upon request for participation. Also, our ethics approvals from Muhimbili University of Health and Allied Sciences and National Institute for Medical Research’s Ethics Committee were granted based on the anonymity of the individuals consenting to participate. Due to these conditions, the authors are unable to avail the full transcripts. Excerpts of specific segments of the text will be reviewed for any potentially identifying details and made available to fellow researchers or reviewers who complete a data sharing agreement and abide by strict confidentiality protocols. In line with the information given to the participants and restrictions set by the ethics committees above, access to the full transcripts is only available to the involved researchers. Data requests may be sent to the corresponding author, RU, via.

## ACKNOWLEDGEMENTS

We are very grateful to all participants from included hospitals who so generously and openly shared their opinions and agreed to be followed during observation. We would like to thank regional, district and hospital managers who allowed us to enter the hospitals and conduct our research. We acknowledge the outstanding work of our colleagues and additional data collectors from Muhimbili University for Health and Allied Sciences Mr. Emmanuel Massawe, Mr. Ruchius Philbert and Mr. Rwegasira John, who also assisted with transcription of the audio files. We are also grateful to Ms Maria Berndtsson for her assistance with the illustrations.

## AUTHOR CONTRIBUTORSHIP

RU developed the study design, contributed to data collection, conducted analysis and drafted the manuscript. FAA contributed to the data collection, analysis and contributed to manuscript writing. ZJ and EM contributed to data collection and analysis and to manuscript writing. ABP and CH contributed to the study design and manuscript writing. HMA contributed to the study design, analysis and manuscript writing.

## COMPETING INTERESTS

All authors declare no competing interest.

## FUNDING

This study is part of the ALERT-project which is funded by the European Commission’s Horizon 2020 (No 847824, PI Claudia Hanson) under a call for Implementation. research for maternal and child health. The contents of this article are solely the responsibility of the authors and do not reflect the views of the European Union (EU). The funder had no role in study design, data collection and analysis, decision to publish, or preparation of the manuscript.

## PATIENT CONSENT AND ETHICAL APPROVAL

Written consent from participants and written institutional consent for observations at maternity wards as well as assent from MCPs, were obtained. Clearance was obtained in Tanzania from the Institutional Ethics Review Board of *Muhimbili University of Health and Allied Sciences* (MUHAS-REC-4-2020-118), from the *National Institute for Medical Research* (NIMR/HQ/R.8a/Vol IX/3493) and from the *Swedish Ethical Review Authority* (2020-01587).

## Notes

### Competing Interest Statement

The authors have declared no competing interest.

### Funding Statement

This study is part of the ALERT-project which is funded by the European Commission’s Horizon 2020 under a call for implementation research for maternal and child health. The contents of this article are solely the responsibility of the authors and do not reflect the views of the European Union. The funder had no role in study design, data collection and analysis, decision to publish, or preparation of the manuscript.

### Author Declarations

IRB of Muhimbili University of Health and Allied Sciences gave ethical approval for this work. Ethics Committee of the National Institute for Medical Research gave ethical approval for this work. Ethical Committee of the Swedish Ethical Review Authority gave ethical approval for this work.

