## Supplementary figures and images for "Understanding maternity care providers’ use of data: A qualitative study in Southern Tanzania"

### Supplementary figure 1

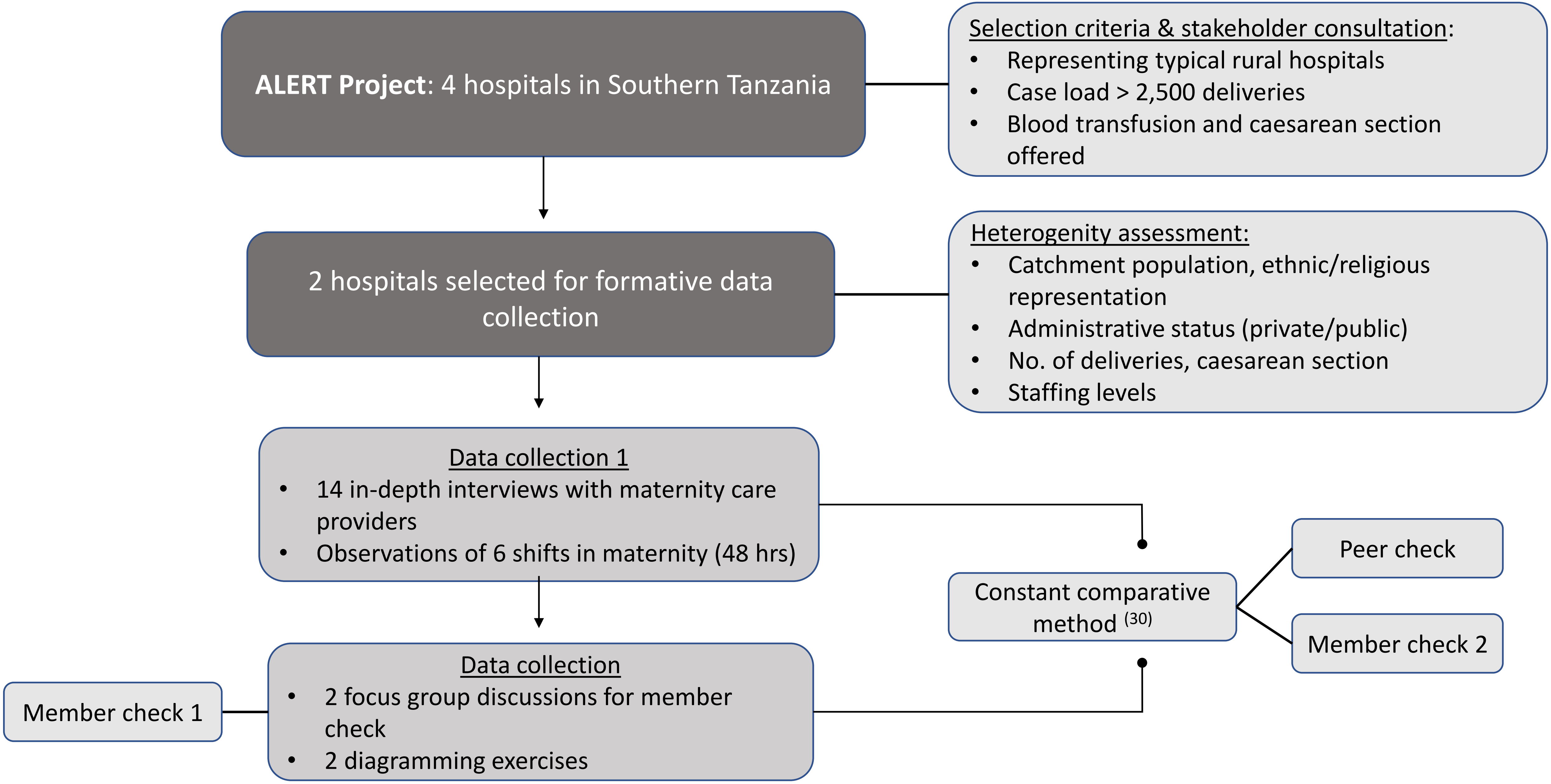
